# Successes and Challenges of an online based nutrition awareness program in 9–11-year-old children In Four Arab Countries: The Ajyal Salima digital platform Qualitative study

**DOI:** 10.1101/2025.05.18.25327873

**Authors:** Carla-Habib Mourad, Carla Maliha, Amira Kassis, Diala Tailfeathers, Marco Bardus, Eman Haji, Lina AlTarazi, Suzan Totah, Nahla Hwalla

**Affiliations:** Department of Nutrition and Food Sciences, Faculty of Agriculture and Food Sciences, American University of Beirut, Beirut, Lebanon; Neat Science, Châtel-Saint-Denis, Fribourg, Switzerland; Department of health and Physical Education, Mount Royal University, 4825 Mount Royal Gate, Calgary, AB, Canada; Department of Applied Health Sciences, School of Health Sciences, College of Medicine and Health, University of Birmingham, UK; Ministry of Health, Building 929, Road 1015, Sanabis 410, Kingdom of Bahrain; Royal Health Awareness Society, Muhammad As-Saeed Al-Batayni St., Amman 11821, Jordan; Ministry of Education, Ramallah 00970, Palestine

## Abstract

**Introduction.:** Digital technologies are increasingly influencing children’s lives, with many seeking digital platforms for nutritional education. This study aims to assess the usability and acceptability of Ajyal Salima, a nutrition awareness digital platform targeting children aged 9-11, in four Arab countries.

**Methods.:** A qualitative study was led across four countries: Lebanon, Bahrain, Palestine, and Jordan. Semi-structured focus groups were conducted with children and parents, and one-to-one interviews were held with teachers. Data was analyzed using thematic content analysis.

**Results.:** Four major themes emerged: platform’s usability, content enjoyment, changes in children’s habits and recommendations to improve the platform. Overall, parents and teachers found the digital experience positive and useful and the content appropriate for children, particularly younger age groups. Challenges included registration difficulties, technical problems, internet accessibility, low parental involvement, and difficulties integrating the platform into teachers’ schedules. The platform’s animations were less effective in sustaining children’s attention amid evolving digital standards.

**Conclusion.:** To enhance the platform’s effectiveness, recommendations include simplifying the registration process, enhancing content interactivity, aligning the platform with school curricula, and equipping teachers with supportive resources. Fostering stronger school-family partnerships and engaging parents through community initiatives may be considered to maximize the platform’s potential to promote healthier eating habits and improve nutritional awareness among children and their families, across the region.

## Introduction

In today’s world, technology is increasingly prevalent, and children. are exposed to digital technology from an early age, with social media being integrated into their lives. Subsequently, there is a growing concern about the impact of media on children, as reported by teachers, parents, and health professionals [1]. Prior to the pandemic, digital interventions were already appealing to young people [2]. However, during COVID-19, the shift to online platforms intensified. Social media usage surged, particularly among youth, as individuals adapted to online learning and engaged more with health-related content. This period saw an expansion of digital health interventions and a growing reliance on social media for health information [3], making digital technologies even more prevalent.

Studies indicate that children and youth often prefer digital methods for learning about nutrition, viewing them as more relevant and impactful compared to traditional approaches, mainly due to their exposure to technology and the digital era [2]. Several platforms are available, including the internet, telehealth, gaming, social media, mobile apps, and wearable devices. Each of these technologies can play a pivotal role in enhancing children’s health outcomes [2] and research from different regions of the world and age groups has shown promising results from digital educational interventions.

Research from different regions and age groups shows promising results from digital educational interventions. In fact, eleven website interventions targeting diet and physical activity (PA), providing nutrition education reported significant improvements in both diet and PA [4]. The KickinNutrition.TV (KNTV) program, an evidence-based digital nutrition and wellness curriculum designed for school-age children, has been shown to improve dietary behaviors and the ability to identify healthy dietary options [5]. This innovative curriculum utilizes digital technology, peer education, and online activities to enhance learning both at home and in school for middle school students. In Greece, the online educational initiative “Nutritional Adventures” has demonstrated effectiveness in increasing adherence to the Mediterranean diet, enhancing fruit and vegetable intake, and promoting PA among children [6].

A systematic review of six articles from different countries examined schoolchildren aged 8 to 16 and various types of games, including computer based, video games and mobile applications. In three studies, children in the intervention group consumed and selected more fruits and vegetables (F&V). Similarly, in two other studies, the intervention group chose and consumed more healthy snacks compared to the control group. One study reported a decrease in sugar intake, although no significant difference was noted between the intervention and control groups [7]. These findings align with those discussed in another systematic review of fifteen studies on website interventions, which highlighted that the success of digital interventions for health improvement depends on multiple factors such as education, goal setting, self-monitoring, and parental involvement [8]. Adding to this, a meta-analysis of twenty randomized controlled trials, involving studies from the US, the United Kingdom (UK), New Zealand (NZ), Canada, and several others, further explored the impact of serious games on children’s health. It examined the effect on body composition, physical activity, and dietary change in children and adolescents. In particular, 10 studies focused on physical activity, involving 489 participants in the intervention group and 425 in the control group, with intervention durations ranging from 1 to 7 months. Most of these studies reported a significant increase in physical activity, highlighting the positive impact of serious games. The dietary change studies, involving 1796 participants in the intervention group and 1913 in the control group, measured snack consumption, F&V intake, and sugar intake. Results indicated a significant reduction in snack and sugar intake, with a statistically significant increase in children’s F&V consumption observed in only two studies [9].

The Ajyal Salima initiative is a school-based nutrition education program in the Middle East and North Africa (MENA) region, designed to raise awareness about healthy nutrition and physical activity in a fun and interactive way. It has been implemented in four Middle Eastern countries over several consecutive years, with recent findings published in a regional paper summarizing its efficacy on different outcomes across these countries [10]. Briefly, the Ajyal Salima program has proven successful in instilling concepts of healthy eating and physical activity in children 9 to 11 years of age and in changing food consumption behaviors and self-efficacy in this age group [10].

During the COVID 19 lockdowns, the need for a digital platform became even more evident, adding more value to the overall initiative and to the future updates of the Ajyal Salima program. The digital platform was developed to extend the program’s reach beyond the traditional school setting, enabling students to access educational materials in the form of modules from home or other locations. The implementation of the educational modules is planned for the academic year, spanning around 3 to 5 months.

The primary objective of this pilot multi-center study was to qualitatively evaluate the usability and perceived value of the platform and identify potential gaps by gathering feedback from students, teachers, and parents, ultimately aiming to optimize its use as a tool to increase nutritional knowledge and improve children’s dietary and Physical activity behaviors. Given the importance of parental involvement in the success of digital education, the Ajyal Salima platform targeted parents by inviting them to take part in their child’s learning.

## Materials and Methods

### Participants

Participants were school children from four Arab countries (Lebanon, Jordan, Palestine and Bahrain), aged between 9 and 11 years and enrolled in grades 4 and 5, along with their parents, classroom teachers and Ajyal Salima staff. The staff included field supervisors from participating countries responsible for conducting regular schools’ visits to follow-up with teachers on the implementation of the intervention. All staff members received research ethics training. The study included schools selected by the Ministries of Health and Education within their respective jurisdictions. Schools were enrolled in the program based on their ability to conduct the intervention as per protocol, including the availability of necessary staff and facilities. The pilot schools were contacted through the Ministry of Education in each country, responsible for approaching schools, obtaining consent, and implementing the program locally. Consent forms providing comprehensive information on the study’s objectives, method, and risks were sent to parents through the schools. Parents signed consent forms for both themselves and their children. Only children with parental consent were approached for participation and all students were required to sign assent forms. Ethical approval of the study was granted by the Institutional Review Board of the American University of Beirut (AUB) in Lebanon (SBS-2021-0423).

### Design and procedures

This qualitative feasibility study was run from May 2023 to May 2024 using focus groups and individual interviews. The intervention was implemented over the course of one month, followed by data collection in all participating countries.

Teachers were introduced to the program and the digital platform through training workshops provided by the research team. While the platform was designed to be used at home, the teachers were instructed to assist children in setting up their accounts and logging in and monitoring their progress throughout the intervention period. Along with the consent letter, parents received a brief overview about the platform including its aim, content and instructions on how to register and log in their children.

### The intervention

The Ajyal Salima digital platform consists of a series of educational modules adapted from the existing offline classroom version [11]. Using an access code, students can progress through each of 10 unique modules on nutrition and physical activity throughout the academic year. The learning modules cover concepts related to food groups and their benefits, portion size, fruits & vegetables, physical activity, importance of breakfast, healthy snacks, importance of water, dental health, sugars and fats hidden in food, and the nutritional value of food. Each module included animated videos and interactive games displayed on the Ajyal Salima Digital platform microsite. The animated videos include two main characters, a snail and a rabbit, who guide students through lessons in a fun and interactive dialogue format. After completing each module, children play a game to apply their newly acquired knowledge. The combination of animation and gamification was designed to make the learning more enjoyable, capturing students’ attention and encouraging them to return to the platform for continued participation.

### Data Collection

After receiving their consent, students were provided with a code to access the online platform, which featured 10 interactive modules to enhance nutritional knowledge and awareness through engaging activities and games. Each code was unique to the teacher and her class, allowing the teacher to monitor and follow up on the progress of the students throughout the modules.

Students accessed the e-educational activities at home at their leisure; all the modules were completed during the intervention period, around one month, but most students completed the whole program within a week or less.

To evaluate the user experience of the digital platform, qualitative data collection was conducted post-intervention (once students had completed all modules) with children, parents, and teachers. Parents and children participated in semi-structured focus group discussions, while teachers were engaged in one-on-one interviews. In addition, the Ajyal Salima staff provided their input on the implementation through regular follow-up meetings and calls.

For children and parents, two semi-structured focus groups per category were conducted, where students and parents had the chance to provide qualitative data about their experience with the Ajyal Salima digital platform. Parents and students who elected to engage in this process were encouraged to share their opinions freely and were assured that all personal identifying information was kept confidential.

For teachers, one-to-one interviews were conducted with the main reference teachers of grades 4 and 5. In one Lebanese school, an interview was conducted with the school nurse, as she served as the school health educator. Each of the interviews and focus group lasted approximately 40 minutes.

Focus groups and interviews were conducted by independent interviewers appointed by each country’s health or education ministry. These interviewers attended 2 training sessions, (1) One on Research Ethics and a tutorial on how to use the Ajyal Salima microsite and two (2) on how to conduct qualitative research.

Regular online meetings were held to collect and synthesize feedback from Ajyal Salima staff across different countries during and after the program’s implementation. A regional debriefing session took place in Istanbul, Turkey, where team members from Lebanon, Jordan, Palestine, and Bahrain gathered to assess the platform’s functionality, discuss implementation challenges, and identify potential solutions to improve user experience. This collaborative meeting enabled staff members to exchange insights and share best practices from their respective countries, fostering collaboration and driving improvements for future developments.

### Data transcription

Focus group discussions and face-to-face interviews were conducted using a set of pre-determined core questions, however they were intentionally designed to encourage open (informal) conversations, allowing participants to express their views freely. Focus groups and interviews were conducted in colloquial Arabic to ensure cultural relevance and clarity for the participants.

To ensure accurate data collection, all sessions were recorded via auditory means to ensure that all information/feedback provided by participants was documented and to preserve the authenticity of the feedback. The recordings were manually transcribed verbatim in Arabic by a designated transcriber in each study country. These transcripts were sent to the research team at the American University of Beirut (AUB), in an anonymous and non-identifiable format to maintain participant confidentiality, for analysis. Transcripts were translated from Arabic to English.

### Data storage and confidentiality

The recordings made during the focus groups did not include names or identifiable information; they only included the questions and answers to the pre-determined interview questions and were not shared with any other party. All soft data were stored on a password-protected laptop, ensuring that only the researchers involved in the study had access to the password and, consequently, the data. As for hard copies/papers (used for documentation of focus group discussion, in Lebanon), they were securely stored in a key-locked room at the department of Nutrition and Food Science and only researchers included in this study had access to the papers. This approach ensured that all data was handled in a way that maintained participant privacy and adhered to ethical standards for confidentiality and data security.

### Data analysis

All transcribed data were analyzed using thematic content analysis, a method described by (Burnard, 1991). Thematic content analysis is adapted from grounded theory and was carried out using a systematic approach of immersion in data, coding, and data reduction. Quotes were inductively organized around four main themes: 1) *Usability and Support using the Digital Platform*; 2) *Content Enjoyment of Story Lines and Games*; 3) *Changes in Children’s habits*; 4) *Recommendations to Improve the Digital Platform*; the mind map in figure 1 (S1 Fig) illustrates the main themes and respective subthemes. NVivo software and established qualitative analysis procedures were used for data review.

**Figure 1:**
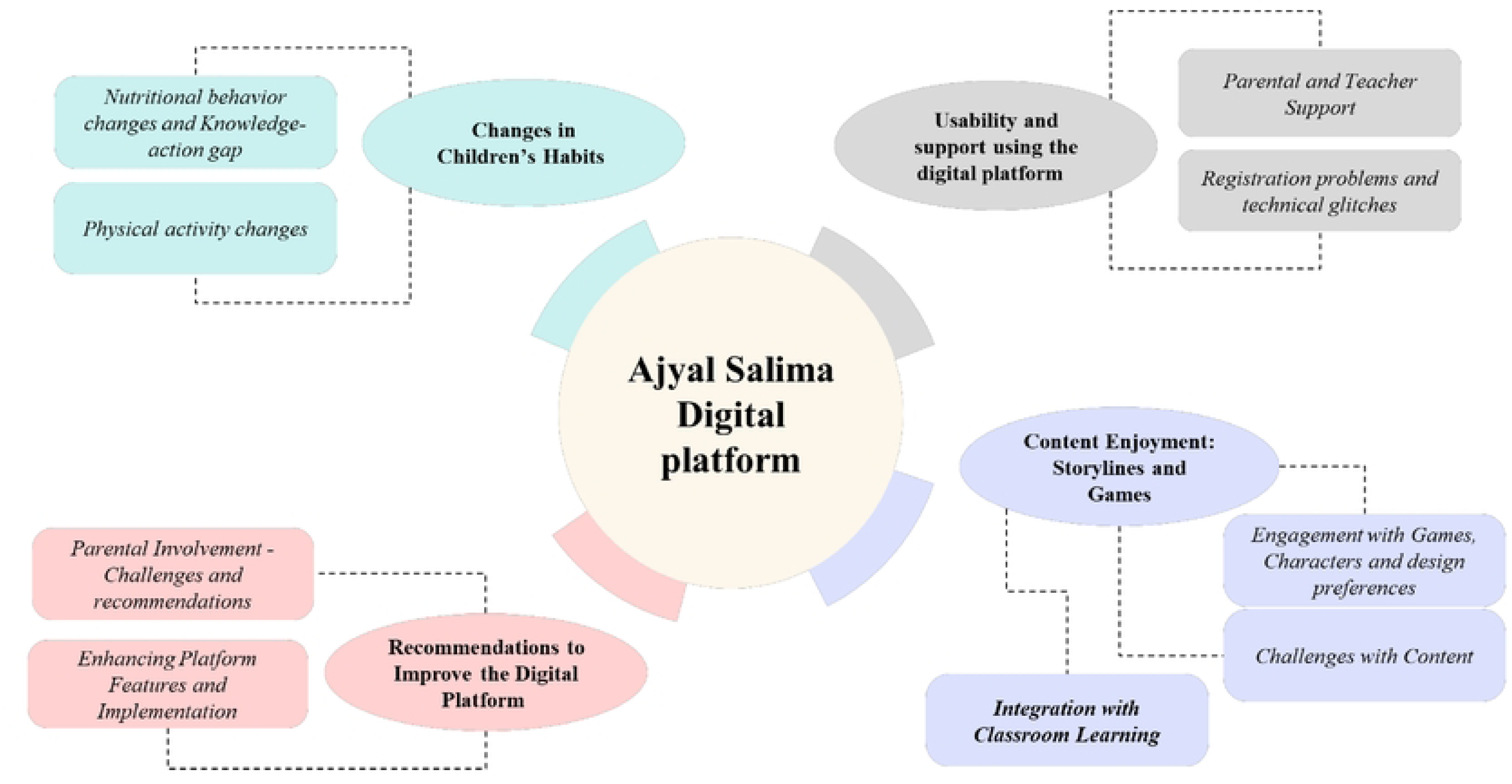
Mind mapping of identified themes and sub-themes.

## Results

Data collection took place in three schools per country, and four schools in Lebanon, for a total of 13 schools. Table 1 details the number and distribution of focus groups, schools per country, and participants.

**Table 1:** Ajyal Salima Digital Platform – Total Number of Focus Groups Conducted and Participants Reached.

|  | **Lebanon** | **Bahrain** | **Palestine** | **Jordan** | **Total** |
| --- | --- | --- | --- | --- | --- |
| **Number of Schools** | 4 | 3 | 3 | 3 | **13** |
| **Number of Children Focus Groups** (Total number of participants) | 7 (43) | 5 (28) | 5 (37) | 4 (37) | **21 (145)** |
| **Number of Parents Focus Groups** (Total number of Participants) | 3 (12) | 4 (20) | 5 (37) | 4 (30) | **16 (98)** |
| **Number of Teachers Interviews** (Total number of participants) | 7 | 4 | 3 (4) | 4 | **18 (19)** |
| **Number of participants from Ajyal Salima Staff** | 3 | 1 | 2 | 2 | **8** |

Themes and their respective sub-themes were extracted, and quotes were organized accordingly. Major quotes with their identified source by country are presented in table 2 (S2 Table). Selected quotes were chosen to support the explanation of the results.

### Theme 1: Usability and support using the digital platform

#### Sub-theme 1.1: Technical Challenges and Registration Issues

Most parents believed that the digital platform was easy to use with clear instructions, that their children needed a bit more time to adjust to the digital platform, but that the overall experience was positive and different from traditional ways of learning. However, some technical problems were encountered such as inserting email addresses, completing the registration process, accessing the link to the nutritional program as reported by many parents, children, and teachers. Collected quotes illustrate this observation, for instance, a parent in Palestine said that *“At first, we faced a problem with the email address where it would get mixed up when we wrote it down, but my husband fixed it for us*” and another in Jordan *“The account login process should be a bit easier than this, not too many accounts and complicated steps.”.* Quotes from students in Lebanon support this issue *“The password was not working”* and *“I got stuck on Step 1 at first—it didn’t work for me, but then it opened, but it was still kind of stuck”*.

A recurring complaint was the glitching of the platform which prevented children from accessing it, interrupting their activities, or even requiring them to repeat activities. This was particularly noticeable in Lebanon, Palestine and Jordan, and was linked to factors such as unstable internet connections, the use of different devices or simply platform related problems. Some examples of these quotes are listed below:

> *“I faced a problem where the platform glitched when I rotated the screen.”*
>
> *“I did not receive the certificate since the seventh activity glitched and I couldn’t proceed. I replayed all the activities to make sure they are all unlocked, and this activity still did not unlock.”*
>
> *“I also have a slow internet connection which makes the platform glitch.”*
>
> *“I did not have any difficulties with the food groups activity… and the activities were easy, but they kept glitching, so I asked for help.”*

These concerns were confirmed during teachers’ focus groups, particularly in relation to technical issues with the digital platform. They reported that many students had difficulties with registration, and needed support to sign in, create passwords, and/or access lesson plans or interactive games. For example, according to one teacher in Jordan, “*The issue was mainly with the registration process from the start. One of the problems was the nickname field”.* Another teacher shows the impact of these difficulties on the actual participation to the program by saying that *“they* (the children*) found it a bit difficult because, honestly, only a small number of students managed to access the platform”.* In Palestine, one teacher said *“The students and their parents also need help since the registration process was a bit of a hassle”*.

#### Sub-theme 1.2: Parental and Teacher Support – Role of teachers and parents in helping children adapt to the platform

Despite the reported issues in registration and some activities, all parents believed that their children received continuous technical support when needed, especially by their teachers, who were, according to parents, always available to help and maintained regular communication regarding the digital platform. This was particularly obvious in the quotes collected in Bahrain: *“They did not need assistance because the teacher was beside them, helping them”,* or *“Yes, there was great support from the teachers, as they even provided us with instructions on how to log into the platform. They also maintained continuous communication with us through contact channels”.* Parents of other countries also mentioned some support from other family members although most of the support came from teachers. In Palestine for instance, a parent mentioned that *“Her (daughter) teacher and older sister helped her”,* while another said *“My daughter had some help from her aunt but was able to proceed on her own later”* However, the teachers complained that the registration process and follow-up was overwhelming given their overloaded schedules *“Time is a conflict since there is already too much to teach and little time to do so.”*

While the registration phase posed some challenges, many students were able to navigate the digital platform independently once registered, as this Jordanian student stated *“Usually, my mom opens the platform for me and I play on my own”.* Another student from Jordan agreed by saying that *“My mother helped me initially with the registration and password, then I was able to play alone since it was easy for me to open the platform and play”.* However, most students mentioned requiring repeated assistance for logging in, or during some activities, be it for content or for technical issues such as glitching. Students generally thought that teachers and parents cooperated to provide support when needed. Many teachers provided follow-up discussions around healthy eating and activities. In addition, punctual or repeated help was requested from parents, usually mothers, however in some cases other members of the family including older siblings were mentioned. For instance, one student in Palestine stated that *“My mom helped me twice in the rainbow activity”* and another student said, *“I needed my mom’s help in the first and last activities, and I have some trouble remembering the information in the sugar and fat activity”.* In Jordan, one student said, *“I found the food groups activity hard at first, but I was able to solve it with the help of my sister”*.

### Theme 2. Content Enjoyment: Storylines and Games

#### Sub-theme 2.1: Engagement with Games, Characters and design preferences

Overall, students liked the platform’s digital format, which included interactive games, videos, pictures, and the characters. They found the platform both entertaining and educational, and several students wanted to involve other friends or siblings to join. This is represented in the following quotes from students in Palestine *“I would recommend it to younger age groups because it is fun and so that they stay away from unhealthy foods. The platform itself is nice and encourages us to play*.”, Jordan *“I would play again but it would be better if you updated it and added more videos and new information that we can benefit from. My younger siblings used to watch some of the easy videos with me so they can learn from them too”,* and Lebanon *“Their voices were funny, and they explain very well. The information was easy.”*

All parents believed that the content was age-appropriate and that the games and videos on the platform were new and attractive to their children. One parent in Palestine thought that *“Everything was suitable for their age group and the characters were nice and entertaining”* In Jordan, parents were also very positive about the appropriateness of the format and content of the platform as can be seen in this quote from one parent *“The information is presented in a simple and nice way using cartoon characters which is nice and suitable for kids”* and another *“The platform is using an approach similar to video games which is attractive to children and makes the information stick to their minds.”* Also, a parent shared that his daughter *“liked all the activities”* specifying that *“What is attractive, is the presence of games, entertainment coupled with great information, and a certificate at the end”*.

Some parents liked the point collection system, believing it encouraged children to participate and stay motivated. One parent from Palestine said, *“She also got excited when she was ranked first and collected points.”* and another confirmed with this quote *“The concept of collecting points was also a great idea that created a competition between the girls.”* This aspect of competition was appreciated by parents in Jordan as is mentioned in this quote from one parent *“I think what attracted the children most to the platform are the things that can include competitions like the games”* and recommended by some parents as further improvement. Notably, in Bahrain one parent said, “*The program should be continuous, with different levels, and the addition of some competition activities”*.

Parents also expressed that the use of vibrant colors and appealing characters as well as the varying difficulty levels in online games contributed to their children’s motivation. One parent from Bahrain said *“The characters were fun and made learning exciting, our favorite character was Fareed the rabbit. The information was easy to understand, and some of it was already covered in the family education subject”* another parent in Palestine mentioned that *“The presence of cartoons and games (especially the baskets game) was fun, and she liked when she got the certificate”.* According to teachers, in the beginning, some students were not particularly excited about the digital platform but became more motivated once they started the games. For example, one teacher in Bahrain said *“It was a new experience, but the students enjoyed it and were excited to complete the activities. It was a good alternative to electronic games.”* That said, they believed that the digital platform provided an effective novel learning opportunity that kept students enthusiastic and interested. According to one of the teachers in Palestine, *“The students enjoyed using the platform and were encouraged to proceed. They were accepting this new idea and were happy with their experience.”* The cartoon characters and interactive games did catch their attention. For example, according to one teacher from Jordan said *“They enjoyed it because of the existing drawings, cartoons and videos. They enjoyed the fact that they can play and collect points, and that there is a competition to collect points from within the class and among the girls.”*

#### Sub-theme 2.2: Challenges with Content – Difficulty with certain lessons and lack of engagement in certain videos

Overall, this theme highlighted differences between countries, with Palestine reporting the most hardship with some activities. Also, the age of children plays a role in the appreciation of the videos and the appropriateness of the digital interface.

Although the content of the platform was considered easy as seen in previous sections, some challenges were reported in the different focus groups. The difficulties mentioned by parents were generally subject-specific. For instance, in Palestine and Bahrain, the food portion activity was deemed difficult as evident from the following quotes *“She had some difficulties with the food portions activity”* or *“The most challenging games were “food portions,” and the “maze dice”.* A common difficulty mentioned by students in Palestine was the lesson on fats, which some found challenging due to the speed at which the information was presented. In fact, eight different children said that the activity on fats was the hardest one, where they needed the most help. These examples of quotes reflect the difficulty experienced by students in Palestine: *“The video where they were talking about the fats and oils was a bit hard.”, “The sugar and fat activity is the hardest.”* And *“All the stories were easy except for the one about fats.”* This was not the case in Lebanon and Jordan where children thought the activities were easy, more suitable for younger kids, and sometimes boring. A student in Jordan said: *“I suggest making the activities a bit harder because I thought they were too easy and students younger than us could do them”*. Similarly, a student in Lebanon stated that *“I felt it was for younger kids”* and another *“Easy but the videos were long”.* Finally, some students, generally older, lost interest in the digital interface. For instance, some grade 5 students found the interactive design less engaging. One student in Lebanon said, *“One of the videos was boring I had to leave the phone and come back to do the activity”* and another *“The activities were ok, but I felt that the videos were better for younger the characters and how they talk are designed for younger, maybe they can add real characters.”* Some students reported that longer videos, those lasting between 4 to 6 minutes or longer, did not hold their attention, leading to disengagement.

One teacher in Lebanon confirmed the lack of motivation of students to use the platform by highlighting the importance of finding different ways of engagement and motivation. No other statements were collected by teachers that reflected the challenges faced by students with the content except for one teacher in Jordan stating that *“The activities that required the students to memorize a lot of new information were the hard ones.”*

#### Sub-theme 2.3: Integration with Classroom Learning – Teachers’ perspectives on how the platform complements traditional teaching

The use of this digital platform in the classroom is viewed to be a valuable complement to the curriculum, a means to break away from the rigidity of traditional teaching and introduce more interactive and flexible learning experiences. Many teachers believed that the content of the platform could be easily integrated into the classroom, more specifically in science class. As one teacher from Palestine explained, *“As a science teacher, I can include the food groups in my classes by using the lessons, videos and games from the platform, in case a computer was available.”* Another teacher from Jordan explained that the platform could fit her current science curriculum as she tackled common subjects, in her quote *“Sure, for the future, I can benefit from the platform in my lessons. I tackle food groups in my classes, like carbohydrates and proteins. As for the fourth grade, there are full lessons about these subjects*”. In Lebanon, teachers also had a positive attitude towards using the platform as a teaching tool as shown in this quote from a teacher: *“Sure especially when explaining the food pyramid, I could use some of the games as a supplemental teaching tool.”* Another teacher in Lebanon valued the practical aspect of the platform compared to conventional learning saying that *“I can benefit a lot from this platform in the classrooms. It’s much easier than how we used to do it before—printing posters, bringing materials, and you know, with the current economic crisis, this feels like a more practical solution”.* Interestingly, in Bahrain, teachers were more proactive having already integrated the program in their curriculum or side projects. For example, one teacher mentioned *“I integrated the Ajyal Salima curriculum into the Healthy Mind in a Healthy Body project, as well as into the Family Curriculum”* and another said, *“The program was discussed and implemented in physical education classes, where students participated in preparing a healthy group breakfast.”*

Although parents did not express their opinions about this topic in most countries, quotes collected in Jordan highlight the parental support for this platform to be used in the classroom. For instance, one parent suggested *“… implementing the platform’s activities in class too by including one session per week dedicated to this.”* A suggestion with which two other parents agreed.

### Theme 3: Changes in Children’s habits

#### Sub-theme 3.1: Nutritional behavior changes and Knowledge-action gap

Parents believed that their children were choosing healthy foods more often and avoiding take-away meals and processed foods. For instance, a parent from Palestine said *“She stopped buying from the school’s cafeteria and started making herself salads and eating more fruits and vegetables. She also bought less foods from the extra group.”* And another stated *“She started eating more fruits and vegetables, cut down on buying junk food, started eating more sandwiches and yogurt.”* Children were adding more food groups into their meals as observed from this quote from Palestine *“She started eating a wider variety of food and including all the food groups in her meals. She also added yogurt and salads to her diet.* Children also showed an increased interest in food preparation or being present in the kitchen during meal cooking. In Jordan for example, one parent reported *“She’s making her own sandwich that included healthy foods like labneh and tomatoes”* while a parent from Palestine said, *“She started making her own lunchbox and got used to eating breakfast, which inspired her sister to do the same.”* The influence of children’s growing knowledge on the rest of the family transpired in Palestine and Jordan with one example of parent quote from Palestine *“She started eating more fruits and vegetables and made all of us cut down on unhealthy foods”.* Similarly, in Jordan, one parent said *“My daughter started forcing us to eat a side salad every day. She began to cut the vegetables herself saying that we need to include salads in our lunch meal. She also started to cut herself some fruits saying that this is healthy instead of eating dinner.”* Other parents went further by considering that their child was acting like a leader or a coach as highlighted in this quote “She *started teaching her older sisters what’s healthy and what’s not… Even told her cousins ‘You’re eating the wrong way.’ Acting like a health coach.”*

Parents also noted that their children became more interested in healthy living in general after participating in the program. One parent in Palestine noticed that her child “*Became more committed to brushing her teeth regularly.”,* a statement that was mirrored by a parent from Jordan saying, *“When she watched the video on the platform, she started brushing her teeth every day”.* The change in interest and awareness was equally noticeable for parents in Jordan for example *“…my son once picked up a box of juice and told me that it contains a lot of sugar and other ingredients that are not healthy.”*

All teachers believed that Ajyal Salima platform contributed to improving nutritional knowledge which reflected positively on students’ dietary habits; this included healthy food preparation and healthier alternatives in lieu of fast food and sugary drinks. For example, according to one of the teachers in Palestine, *“I noticed an increase in their consumption of healthy foods like fruits and vegetables since they focused on getting 5 servings per day”.* Similarly in Bahrain, a teacher stated that *“Students’ nutritional habits and concepts have changed. They became more consistent with having breakfast, avoided fast food, soft drinks, and consuming from the extras groups….”* In Jordan, 2 teachers had similar positive comments regarding the implementation of the children’s learnings saying that *“They even started implementing the things they learned from the platform at school by brining healthy lunchboxes and showing them to me.”* and *“They started bringing healthy sandwiches and healthy snacks with them to school, and cutting down on sugar, chips, chocolate and juices. In addition, they started drinking more water and playing sports. I noticed these changes since they would come up to me during class and show me how they are drinking more water.”*

From a student’s perspective, all agreed that the digital platform was diverse and that all activities were useful for their learning. They enjoyed learning about healthy fats and new ways of making healthy choices. Students showed noticeable changes in their dietary habits, including reduced sugar intake, fewer snacks, better portion control and increased physical activity. They were aware of their own changes and related them directly to what they had learnt on the platform. For example, one child from Palestine said *“I started drinking more water and eating more fruit and vegetables. For instance, I ate 4 carrots yesterday because it was mentioned on the platform”* while another stated *“I benefited from everything on the platform. I started eating more fruits and vegetables and less from the extras”.* This was also seen in Jordan and Lebanon:

> *“I prepared a diet schedule on a paper that I hung in the kitchen and that helped me know how to eat at lunch. I also used to eat a lot of food that are high in fats and sugar instead of having healthy snacks or breakfast, but after using the platform, I sticked to the diet schedule I prepared and started eating healthy foods”.*
>
> *“I used to eat a lot of sweets, but after watching the video and doing the activities, I learned a lot.” “Breakfast—when they told us what to make for breakfast and how to prepare the sandwich, I liked it and kept making it!”.*

Although student quotes from Bahrain on this particular sub-theme were scarce, two quotes collected during focus groups with students align with quotes from other countries:

*“Changes occurred in our understanding and dietary habits. We reduced fast food and soft drinks, became more consistent with healthy eating” and “We changed a lot, and you can see it in our daily routine. We committed to having breakfast, eating more fruits and vegetables, avoiding soft drinks, and playing sports.”* Finally, students who completed the program seemed to have more knowledge and awareness around healthy eating and are believed to influence their peers in making healthy choices. Another teacher in Palestine explained *“Their knowledge increased, and we did a revision of the food groups. As for their food intake, they cut down a bit on the foods belonging to the extra food group”* and in Jordan, one teacher alluded to positive peer competition in her statement: *“They would also tell me how they got jealous from each other and started implementing the same lifestyle changes. For example, they started buying healthier items from the school’s cafeteria. The students understood the material and started shifting towards a healthier lifestyle, which was also reported by some parents. Some students started eating better and building healthier habits.”*

Despite the positive feedbacks from parents and teachers, some statements collected in focus groups disagreed with the above. Indeed, a few teachers stated that while knowledge and awareness around healthy eating increased, actual implementation did not. This was recorded in Jordan where two different teachers emphasized the contrast between the knowledge and its application in the following quotes: According to two teachers in Jordan, no major differences were noticed in children as noted in these quotes *“No, they are still buying chips and chocolate. They are aware and understand, but in terms of application they did not, not even when I gave them the food lesson”* and “*Personally, I did not notice any changes, but I understood that they retained the information they acquired from the platform”.* Similarly, in Palestine, a couple of parents reported not seeing any changes in their children’s habits, despite better knowledge. For example, one parent said *“Her knowledge increased but her behavior did not change”* or another insinuated that the progress is slow and insufficient by saying *“We are still learning… but the implementation is slow… She cut down on drinking sodas to only once per week.”*

#### Sub-theme 3.2: Physical activity changes

All participants agreed that their kids’ physical activity levels improved after using the digital platform. In fact, many statements reported that children started exercising more often, playing sports daily, jumping rope, using a bicycle, or following the sports video routines. Parents reported increased energy, daily movement, and enthusiasm for sports. For instance, in Palestine, one parent stated, *“My daughter started being more physically active and became more energetic”* while another parent said, speaking of her daughter, *“She wants to play more sports willingly, which felt like an obligation for her before using the platform”.* In Jordan, when asked if they had noticed any changes, one parent in Jordan said *“Ah, a lot, a lot, especially sports. From the day she saw the sports (activity), she got excited.”* Another commented on how the sports activity was her daughter’s favorite *“The sports activity. My daughter started playing with her sister, who’s in the seventh grade, and another girl. They would open the sports activity and do it together.”*

Children mentioned explicitly that they started exercising more and even at specific times like morning routines. They explained that they now included physical activity in their daily schedule and felt more energized. For example, a student from Jordan said *“I started doing more physical activity since I used to be more sedentary. Watching the videos motivated me to be active and taught me that sports are extremely beneficial to our body.”* Another student stated that *“Before using the platform, I didn’t do any kind of sports. But after using it, I started playing sports with my siblings.”* Similarly in Palestine, children reported an increase in physical activity saying *“I started playing more sports”*, or *“I started exercising more”*. In Bahrain and Lebanon, although the collected quotes were limited, they were aligned with those collected in Jordan and Palestine.

The sports activity was for some students an opportunity to share a moment of play with siblings or friends, for example one parent from Palestine said that her daughter *“Started doing Zumba dances with her sister… ‘I am the hero’ game”* and another in Jordan said that *“The activity I remember the most is the sports activity since my daughter would put it on and start doing the moves. She also started moving more instead of sitting all the time.”* This was confirmed by a couple of student quotes from the same countries.

### Theme 4: Recommendations to Improve the Digital Platform

This theme provided valuable insights, offering critical feedback for the research team on areas for improving the digital platform. The recommendation themes listed below cover issues beyond technical improvements where some recommendations were already mentioned in theme 1.

#### Sub-theme 4.1: Enhancing Platform Features and implementation

Parents provided positive feedback on the digital platform and also suggested some recommendations to improve its effectiveness. These included expanding accessibility of the platform to more age groups as explained in this quote from a parent from Palestine, *“I suggest developing the platform to make it accessible for all age groups”* and two other parents from Jordan who said *“I suggest introducing the platform to even younger age groups so they can learn at a younger age how to lead a healthy lifestyle”* and *“I would suggest introducing the platform to even younger age group since they will be able to absorb the information too. It is good for our kids to get used to eating healthy foods instead of junk foods at an early age”*.

Parents recommended to include sharing features allowing children to share photos, videos or progress on activities but also the application of their learnings at home. Some parents even suggested setting up a WhatsApp group for parents to support their children’s progress. For example, one parent from Jordan suggested *“The students can also share their own experience on the platform itself. I suggest also creating a WhatsApp group at the beginning of the school year that includes some mothers who can supervise the progress on the platform. For example, they can do a competition during each semester where mothers can log what activities her children did each day.”* In Palestine, one parent said, *“I suggest adding a summary of the activities at the end either through pictures, voice recording or educational tool, as well as adding an option where students can upload pictures and videos of the activities they do at home”.* To which three other parents agreed. Parents also concurred that gamification and competition could further motivate their children. By adding points, rewards, and rankings, parents believed that their children’s motivation would be increased. For example, a parent in Bahrain stated, *“The platform needs challenge-based activities between two people to increase excitement, along with (notes) to allow self-reflection.”* Although facilitating the registration process was quite a frequent suggestion, the most common recommendation from parents was to improve and customize characters and make the platform accessible for younger age groups, the quotes from which have been covered above. These two common parent recommendations are in alignment with the students’ suggestions, further emphasizing their relevance. This view was reinforced by students. The two most common suggestions were to add more characters and enable customization of characters, and to add more videos and games. Some notable quotes include*: “I wish they added one more character to the videos”* from a child in Palestine, and *“…We can choose the characters ourselves and customize their clothing, name and even characters where we can choose the animals we like”* from a child in Jordan. Other children suggested that more videos and games be added to enhance learning as was seen in these quotes from two different children in Palestine *“I wish they would give us something new to learn”* and *“I suggest adding more videos so we can learn about health…”*.

As for teachers, they proposed some ways of enhancing the learning potential of the platform such as this teacher in Palestine who said *“I wish there was a practical activity in between games because I felt that the overall picture wasn’t clear”* or a parent from Jordan who suggested *“…. adding a section on the platform dedicated to the takeaway messages that the children can write down after finishing the activities to see what are the things they benefited from. Another thing that can be added is a section at the end in which the child has to do a project or maybe draw a picture that depicts how they benefited from the platform.”*

Students shared a range of ideas focused mainly on improving engagement, content variety, and making the platform more fun and personalized.

One meaningful suggestion that aligns with recommendations made by parents is to add more games, levels, rewards, and interactive features. For example, in Bahrain one child said, *“We also need more motivation, additional levels, more points, and increased opportunities for two-player challenges.”* And in Palestine, two different students stated, *“I suggest adding more games” and “I suggest adding extra levels”*.

Improvement of cartoon characters was a recurring theme for parents, teachers and children. All participants suggested introducing new characters as students’ progress through the platform to maintain interest and introduce variety. In addition, some suggestions were made to customize characters through different clothing, names etc… For example, a parent in Palestine said that *“I suggest adding more colors and characters”.* The same was noted in Jordan with this other parent quote *“…Plus, the characters should change at each stage, so each level has different characters.”* In Bahrain, a teacher suggested *“adding more characters to engage children further.”* And in Lebanon, one teacher’s opinion about the interface was that it needed to be enhanced on the digital side to remain interesting for the children. The teacher said *“In my opinion the videos and activities are neutral since there are much more sophisticated and complicated ideas online”.* Some suggestions were also put forth by parents to develop character-driven stories, turning each learning unit into an episodic narrative to make the platform feel like an ongoing journey. A notable quote from a parent in Palestine illustrates this *“I also suggest creating characters for a specific story to make it more like different episodes in a way that each unit has its own purpose. It’s like creating a 10 episodes series for example with a united background story by changing the way the story is told”.* Finally, one suggestion from a parent in Palestine was to make characters more relatable by changing to a real-life character format. Children also agreed that they would like to see more characters added to the videos and for characters to be more engaging. For instance, in Bahrain, one child said *“we need more engaging characters and additional levels”.* Some children also expressed their wish to be introduced to more recipes *“I suggest adding a section in the platform where we can learn how to make healthy versions of foods, like chips for example*.

Finally, a common recommendation, especially amongst teachers, is to change the timing of implementation, deemed inappropriate as it is too late in the year. In Lebanon, one teacher recommended to start the program earlier, as mentioned in this quote: *“It would be better to implement the program earlier in the academic year because, by the end of the school year, students tend to lose motivation and energy.”* Other quotes from teachers in Palestine and Bahrain confirm that the timing of implementation was not ideal. In Palestine, one teacher said “*The implementation time happened to be during the exams period and the beginning of Ramadan so there wasn’t a lot of time… It would be better if it can be implemented at the beginning of the school year”.* In Bahrain, another teacher stated *“The timing was inconvenient for teachers as it coincided with the second semester, the beginning of Ramadan, and school holidays.”*

The length of implementation was also a subject of a couple of recommendations from teachers but also parents and students, wishing the program was implemented during the whole school year as some students finished all the activities within a week or less. Both parents and teachers believed that students would have benefited more if the period of implementation had been longer.

#### Sub-theme 4.2: Parental Involvement - Challenges and recommendations

Although parental involvement did not emerge as a meaningful theme in the recommendation theme, discussions in focus groups highlighted challenges and barriers in engaging parents to support their children during the implementation of the platform. To these challenges, some recommendations were offered by parents and teachers. In Jordan for instance, one parent stated, *“The platform is nice but needs follow up and commitment from parents”,* while a teacher suggested to raise parental awareness in her statement *“What can maybe help is to raise awareness among parents. I mean the parents must know whether it is good or bad to pack a chocolate sandwich in their kid’s lunchbox.”* Another teacher in Lebanon went further to suggest how to raise awareness of the platform by proposing *“An open house for parents (for example for an hour) to introduce them to the platform and show them what the program is about”*.

The barriers mentioned by teachers in the 4 different countries are a testament to the need to improve parental involvement for the success of the platform.

Parents were generally not very involved or engaged in their children’s use of the platform. Many of them admitted to either not using the platform themselves or not following up on their children’s activities. For example, a parent from Palestine said *“To be honest, I did not follow up much on the platform…”* or in Lebanon, one parent said *“No, I didn’t notice the platform, but my son went in and tried, and he started making food. But he didn’t tell me anything—I just saw him making food, but I didn’t understand why.”.* The teachers noted this parental absence as can be seen in their quotes, here from a teacher in Jordan saying “*They weren’t much interested in it. Maybe because it is not part of the curriculum or because it happened to be implemented at the end of the school year. I didn’t even feel that their knowledge of the topics increased, they told me that the information is close to what I had already given in class”.* Similarly, in Lebanon, teachers reported that parents were not following up with their children’s progress. This is reflected in the following quotes from two different teachers:

> *“The parents were not involved in the platform and did not follow up on the activities.” and “I was shocked that the parents did not follow-up with their children and did not even check what they are doing on the platform”.*
>
> *“Difficulties faced were mainly in the registration process, especially that parents did not help their children in the process.”*

Although not many children quotes were collected, they confirmed the low engagement of their parents and their lack of support. More importantly, this lack of support was sometimes a barrier to their participation in the program. this was particularly noticeable in the following respective quotes from two students in Jordan: *“I did not do the shoebox activity since my mother was busy and I couldn’t do it on my own”* and *“I couldn’t try any recipe because my mother doesn’t allow me to access the kitchen and did not let me try any recipe”.* In Lebanon and Palestine, similar barriers were encountered as seen in these respective quotes *“Every time I ask my mom something, she tells me, later, I’m busy right now”* and *“There was a lot of information given at once, and no one helped me at home”*.

### Feedback collected from Ajyal Salima Staff

Insights from Ajyal Salima Staff meetings highlighted the various challenges encountered during the implementation of the digital platform. First, the registration process posed the greatest obstacle to accessing the platform. This issue was compounded by various technical difficulties encountered by students, parents, and teachers, limiting the number of participants and their level of engagement. Access to the internet for example was a crucial issue where some families were limited by internet access or internet usage in their homes. Despite technical issues, many teachers and parents shared their content with the digital platform once these challenges were resolved. Initially, the platform aimed to target parents, but unfortunately, there was notably low participation from parents with the platform activities. This could be attributed to several factors, such as time constraints, competing activities with their children, or insufficient access to tablets or phones for their kids to use. Low parental involvement can also be due to the fact that the program was being implemented in schools, which made the parents rely on the teachers and schools to constantly follow up on their children’s progress. Many teachers suggested parent meetings as an important step to describe the objectives of the intervention and expectations around parents’ involvement.

Teachers complained about overwhelming schedules, which hindered them from keeping track of and constantly following up on children’s progress. They also faced issues persuading kids to register, engage in activities, and learn through the platform. In concordance, convincing parents to register and monitor their children’s performance and activities was challenging.

Several other barriers hindered the proper engagement of young users. One significant challenge is the increasing difficulty of creating animations that meet children’s evolving expectations and capture their attention. As children are increasingly exposed to sophisticated augmented reality games, simple or basic animations may fail to engage them effectively.

### Notable Quotes

> *“Parent meetings are crucial for explaining the type of intervention taking place, how they should be involved, and how to use the platform. Informing them through a letter or relying on kids to inform them has not been effective.”*
>
> *“Some parents asked their kids if the platform needed internet, a mom asked me if it needs internet.” Maybe they were worried about the internet data usage.”*
>
> *“Internet connection was always an issue.”*
>
> *“A mom told me I don’t want to give them my phone, I don’t want them to use my phone, I need it” “Some parents understood the overall project but did not follow their children progress or engage with them in the activities. They received information from their kids, saw them doing some activities, and that was it.”*
>
> *“The platform is an additional asset to the program, once the registration issues are resolved”*
>
> *“If the platform was available as an application accessible to everyone, it would simplify the process of logging in and broadening its reach. This way, there would be no need for a school code or limitations on age groups and beneficiaries.”*
>
> *“Some teachers faced issues with the program because they were overloaded and had to prioritize the curriculum over activity programs.”*

## Discussion

The Ajyal Salima platform, designed to enhance dietary and physical activity practices, is unique and comprehensive, addressing both children and their immediate environment with interactive content specifically designed for children. Not only does the platform engage children in educational activities, but it also provides tips for parents on healthy eating habits and physical activity and offers teachers means to engage with students through activities related to healthy eating.

The findings from this qualitative study showed that in general, parents believed the digital experience of the Ajyal Salima program was positive and useful, and that the content was age-appropriate and generally attractive to children, particularly in younger age groups. Although the findings are similar for teachers who also thought that the platform was easy to use and that students were well-assisted, technical difficulties in registering and starting the program were noted and this was a major challenge in the study. These included registration issues, internet connectivity and access to games. Technical problems, which emerged as a primary sub-theme in our analysis, highlighted a certain variability between countries and the need to assess the reality of technological literacy and availability in the studied population. Usability and user experience of a digital educational platform is of utmost importance to ensure that the goal of the platform is reached and that users reap the benefits of the platform. This is evident in the literature which shows that positive correlations are found between usability and improved outcomes and engagement [12–14] be it in the field of health behaviors, creativity, or skill acquisition. A review of 610 studies found that usability evaluation methods directly influence the success of digital health tools, with user-involvement and tailored approaches improving outcomes [15]. Usability inevitably depends on digital or eHealth literacy, an essential determinant or moderating factor of digital health interventions in general [16]. eHealth literacy, defined as the ability to seek, find, understand, and appraise health information from electronic sources and apply the knowledge gained to addressing or solving a health problem [17], is crucial in the digital age. Individuals with higher eHealth literacy are better equipped to navigate online health information, leading to more informed health decisions. Conversely, those with lower eHealth literacy may struggle to discern credible information, potentially impacting their health outcomes [18]. In this case, the feasibility of the Ajyal Salima program highly depended on the availability of connected devices in households, as well as access to and quality of the internet connection and on the ability of the users to make use of the platform. These features are in turn highly dependent on socioeconomic status. It is our recommendation that the digital Ajyal Salima program be adapted to or take into consideration these factors.

Children found the content helpful, appreciating its playful format which promoted collaboration between students and teachers/parents. On the other hand, children recommended better user support to assist them through the program, and some older children found the animations boring. While the snail and the rabbit can be fun and interactive for the lower age groups, older children are nowadays exposed to a multitude of digital tools and games which exacerbate the increasing difficulty of creating animations that meet their evolving expectations, capture their attention, and engage them effectively [19–20]. Therefore, it is essential to engage children during the development phase to ensure that the content resonates with the target age group. This recommendation helps identify areas for improvement and better align the platform with the preferences and needs of young users such as the appropriateness of character design, static backgrounds, and the optimal length of videos.

Although the main aim of the study was not to evaluate the usefulness of the digital platform on nutrition and physical activity habits, interesting insight on the perceived usefulness of the platform was extracted from both student and parent focus groups. Most students believed that their sugar intake was lower, so were their snack intake and portion sizes, while they reported increasing their physical activity. Parents generally agreed with these reported trends, stating that their children were more likely to engage in physical activity and active play, also more likely to avoid high energy dense meals and processed food. Our findings are aligned with the literature on school-based educational programs, showing that such programs can improve dietary habits and nutrition knowledge in schoolchildren [21–23]. In a previous report on the Ajyal Salima offline program, we also reported an increase in healthy eating and physical activity habits, measured through standardized questionnaires [10]. Our preliminary results reported here suggest that the digital platform could be effective in implementing behavioral changes, or at least that the platform can serve as a good vector to convey a message of nutrition and physical activity to schoolchildren. The impact of the digital platform on eating behavior and physical activity should be measured in an intervention trial measuring the benefits perceived in this qualitative feasibility study.

One of the limitations noted in this study is the duration of the exposure to the digital platform. Indeed, although the platform was meant to be used for one month, the access was given such that some children unexpectedly completed all modules within one week. While this may show interest in the content of the platform, it also calls for increasing the amount of material in each module or restricting daily access. The literature on digital educational platforms is broad and spans across different disciplines, however it suggests that these platforms are more likely to show measurable behavioral changes in schoolchildren when used consistently over periods of 3–6 months, especially when they target nutrition and physical activity behavioral changes [24–25]. This is supported by a systematic review on digital intervention strategies for diet and physical activity concluding that several weeks to months were necessary for these interventions to be effective in changing health-related behaviors. Indeed, research shows that short-term interventions have limited impact, and sustained engagement is key for the success of digital educational initiatives [8]. Engagement is a fundamental dimension in behavioral programs and digital health apps for weight management, nutrition and physical activity promotion [26–27]. Sustained engagement, combined with features like interactive feedback and parental involvement, is also essential for efficacy. Sustained engagement of children is generally promoted by goal setting, self-monitoring within the duration of exposure, and parental involvement, contributing to obtaining the best results in terms of behavioral change [8]. The goal setting aspect of engagement was highlighted in our study, especially in quotes collected from parents saluting the point system and other features that fostered competition amongst peers. According to parents, these features should be further developed as they provided motivation to learn through the platform.

An important aspect of the platform is that some activities were designed to involve children and their parents. In this context, parental involvement, or lack thereof, can be highlighted as an improvement need and a potentially important determining factor for the success of the educational platform, even in the case of technical feasibility. Indeed, there was notably low participation from parents with the platform activities, and this is potentially due to logistical issues as highlighted during staff discussions. The reliance on teachers for the implementation and follow-up of the program may have also contributed to low parental engagement. Parental involvement is key to behavioral interventions in children as was shown in previous research [28–30], in the public school system and regardless of socioeconomic statuses [28,31]. This was also noted by teachers in a previous paper published by our group on the Ajyal Salima offline program [10]. The 2015 Global School Health Survey conducted in Oman, which focused on parental involvement and adolescent well-being showed a significant correlation between positive parental involvement and nutrition and exercise. Moreover, higher parental involvement was associated with higher odds of good nutrition. Larger studies from the United States and Eastern Mediterranean Region indicate that higher family functioning is linked to better nutrition habits [32]. Another study on Australian adolescents further supports these results, highlighting that maternal knowledge of healthy food items and their availability at home, is related with higher intake of fruits and vegetables and lower consumption of energy-dense snacks [32]. These findings suggest that increased parental involvement is associated with better physical health (i.e., improved nutrition and physical activity) [33].

The impact of parental engagement is far beyond the raising of the child’s interest in the platform. It affects children learning and teacher involvement. This impact was actually reflected in the reduced usability of the platform. A lack of parental engagement and support at home with technical difficulties can lead to disengagement of children, consequently impacting nutrition and PA learning, knowledge, and behavior. There is existing literature underlining a decline in family engagement in both school-based and home-based activities over time, particularly in low socioeconomic communities and amongst individuals from diverse cultural and linguistic backgrounds. For instance, some immigrant families perceive school as an external authority, separating the role of the family in education from the school’s role. Consequently, these families will be less prone to engage in their children’s education at home [34]. In these contexts, parents usually lack the knowledge, confidence, or access to sophisticated technological advancements to support their children’s learning [35–37]. Poor communication between families and schools is a significant barrier to family engagement, discouraging parents from becoming actively involved in their children’s education [35]. Other hurdles include low self-esteem, language and cultural differences, and parents’ tendency to rely on schools, viewing teachers as the experts who are more equipped to educate their children. Adding to these, low literacy levels, unfamiliar educational terminology, and a dislike for meetings [34]. Narrowing it down to the Arab world, eight barriers to parental involvement were identified: time restrictions, student attitudes, poor communication skills, student performance level, parents’ characteristics, parents’ attitudes, and unappealing activities [34].

A lack of parental support was also noted as a reason for the reliance on teachers’ support and potentially causing the teachers’ work overload mentioned by some during the focus groups and the staff feedback meetings. In fact, previous qualitative research exploring the use of digital platforms for behavioral education in children report that the success of these programs is determined by multiple important factors including 1) leadership and organizational commitment in participating schools, allowing for resource prioritization and pedagogical alignment favoring the attainment of the program objective [38] and 2) sound development of teachers and proper training for parents. Indeed, teachers’ ability to experiment and have time allocated to train on the new digital tool could have a strong impact on the success of its implementation [39]. Having time in the curriculum assigned to the digital platform highly depends on the school management team reorganizing schedules to accommodate the implementation of the digital initiative. This time allocation would allow teachers to properly integrate sessions dedicated to digital learning to follow up on the progress of children, solve issues they are facing, and motivate them to keep learning. Time allocation would also enable them to integrate the platform in the current curriculum as noted as a common suggestion by teachers and parents in this study.

Given our observations of low parental engagement, reliance on teacher support and their perceived impact on the use and effectiveness of the platform, we believe that finding the right balance in support between parental and teacher support to students, may be one important improvement to bring to the program.

While it may be challenging to effectively target parents and increase their engagement, the convergence of research findings emphasizing the critical role of parents warrants special attention and suggests that a preliminary qualitative study or a survey of participating parents may be useful to identify ways to engage them. For instance, a Polish study aiming to identify optimal ways to engage parents in their children’s nutritional education program reports that parents are likely to engage with nutrition education platforms that offer practical, accessible content, to support their children’s diet [40]. Therefore, one recommendation for future studies testing a digital platform for nutritional and health behavioral change would be to ensure parental involvement through understanding parental needs and perceptions and identifying ways that best target parents based on their inputs.

Having identified the best way to reach parents depending on their needs and constraints, it becomes easier to establish a sustainable, ongoing practice of engagement. This should go beyond occasional events and involve sustained commitment and collaboration between schools and families throughout the entire school year, rather than just once or twice [35]. In addition, offering parents the option to attend school meetings or webinars online, at their convenience, with guidance and facilitation from teachers, may be an effective way to involve and empower parents and create a stronger alliance between parents, schools and teachers [41].

### Strengths and Limitations

The implementation of the program during this phase demonstrated several strengths, notably the collaboration with local partners and the successful engagement of schools, which facilitated access to the digital platform. The platform’s interactive format and educational content were generally well-received, especially by younger children, and supported the delivery of key nutrition and physical activity messages. Moreover, the platform proved to be a potential educational tool not only for children but also for teachers and parents, offering opportunities for reinforcing nutrition messages through engaging digital content. However, several limitations were identified, the most important ones being the length of the intervention and parental engagement channels. The duration of the intervention may have been insufficient to capture feedback on the usability of the platform, and eventually to influence long-term behavior change. As for parental engagement, parents were primarily informed through a letter sent by schools with no direct interaction. In person follow-up meetings during study implementation may be considered to increase parental understanding and investment in the study. Finally, data transcripts were not subject to back translation, however, the forward translation was reviewed by bilingual researchers to ensure cultural relevance of the translated content.

## Conclusion

In conclusion, the results of our study support the usefulness and usability of this digital platform for nutrition and physical activity education, especially that it fulfills the need for children to learn in a playful way. In this paper, we highlight the challenges and limitations of the intervention in special developing countries’ settings and provide recommendations to improve the implementation of this or similar educational platforms. Our first recommendation is to better assess and understand the socioeconomic reality and digital literacy of households. We also recommend fostering effective partnerships between families and schools as they are essential to ensure supportive milieus that encourage parents to feel appreciated, included, and actively engaged in their children’s education. Moreover, it is recommended to extend the platform’s reach beyond the school setting and actively involve parents, large community campaigns in collaboration with local governments and media outlets. Additionally, children’s engagement can be improved by facilitating the registration process and adapting the storyline and animations and engaging features to their digital experience expectations. And finally, allow teachers time and resources to learn, experiment, and support children in their educational journey. Given the interest shown by teachers in implementing this platform as a classroom hands-on learning tool, this opportunity should be explored to optimize student reach and effectiveness of the platform.

By addressing these recommendations, the platform can better fulfill its goal of promoting healthy eating and PA habits among children and their families. Engaging parents directly on the one hand and teachers through classroom implementation on the other, may create a more comprehensive approach to nutrition and PA education, ultimately contributing to improved health outcomes for children.

## Data Availability

All relevant data are within the manuscript and its Supporting Information files.

## Supporting information

S1 Figure 1. Mind mapping of identified themes and sub-themes

S2 Table 2. Quotes from Parents, Teachers, and Students Categorized by Themes and Sub-Themes

## Authorship

C.H.M. is the co-principal investigator and was responsible for the conceptualization of the study objectives and methodology, has overseen the data collection, digital platform development and the write-up of the manuscript. C.M. contributed to the design of the study, development of the digital platform, and manuscript write-up. A.K. drafted the manuscript. D.A. and A.K. guided the statistical analyses. M.B., contributed to the write-up of the results. C.M., E.H., L.A. and S.T. oversaw data collection in their respective countries. N.H. is the principal investigator, she contributed to the conception of the study and helped with the interpretation of the data. All authors read, provided revisions and approved the final manuscript.

## Acknowledgments

We thank the Ministries of Education in Lebanon and Palestine, Ministries of Health and Education in Jordan and Bahrain; the schoolchildren, their parents and their teachers; The Royal Health Awareness Society in Jordan for data collection and securing approvals; Dina Mansour and Anna-Maria Dannaoui for referencing and proof reading. We also thank Sonia Najem, Hilda Khoury and Fadi Yarak from Lebanon; Reem Jarrar, Dr. Samar Batarseh and Khitam Hattar from Jordan; Nisreen Khaleel Rimawi, Furat Muntaser Ateeq, Lubna Daoud Awwad, Maysa Khalid Al Younis, Sojoud Fawzi Jabra, Sanaa Othman Qawasmi and Aisha Deeb Ghazawi from Palestine; Dr. Mariam Ebrahim AL Hajeri, Dr. Ashwaq Abdulla Sabt, Hessa Khalid ALshaik, Aysha Mohamed Salim, Feda Ahmed AbdulRasool, Lulwah AbdulAziz AlThakir, Saad Eid Saleh, Ahmed Saad Ali, Hameeda Foad AlMudaweb and Enas Salem AlKhalaqi from Bahrain.

